# Evaluating the applicability of replication success metrics in animal-to-human translation: A simulation study

**DOI:** 10.1101/2025.11.07.25339757

**Authors:** Carolyne Jie Huang, Samuel Pawel, Kimberley Elaine Wever, Benjamin Victor Ineichen, Rachel Heyard

## Abstract

Translation failure, in which promising animal study results can not be reproduced in human trials, is a challenge in biomedical research. Metrics for replication success are widely used to evaluate reproducibility, i.e., the extent to which the results of a study agree with those of replication studies. The relevance of these metrics in assessing animal-to-human translation success (or failure) is unclear. We conducted a simulation study to examine whether these metrics can quantify translation success and how their performance varies under different conditions. Using parameters from a meta-analysis on prenatal amino acid supplementation and maternal blood pressure, we simulated animal and human studies under 648 scenarios, varying effect sizes, heterogeneity, animal sample sizes and number of pooled animal studies. Nine metrics were assessed, namely the two-trials rule, meta-analysis, replication Bayes factor, unweighted and weighted Edgington’s methods, golden sceptical *p*-value and three versions of controlled sceptical *p*-value. Most metrics, except meta-analysis and replication Bayes factor, controlled false positive rates under no heterogeneity, but became liberal as heterogeneity increased, particularly between human studies. Translation power (i.e., the probability of true positive translation success) was constrained by the weaker evidence of the two findings; e.g., small sample size in the animal studies resulted in lower translation power. The metric based on meta-analysis frequently indicated success when either of the species found strong evidence, while sceptical *p*-values were more conservative. The sceptical *p*-value that controls overall type-one error and the weighted version of Edgington’s method performed relatively consistently across scenarios. However, no metric was uniformly optimal. Metrics developed for replication studies can inform assessments of translation, but their utility depends on the underlying evidence and assumptions. Using multiple metrics in combination, with attention to their strengths and limitations, is recommended for evaluating the translation of animal findings to human outcomes.

## 1 Introduction

In many fields of medical research, drugs that show promising results in preclinical studies in animals frequently fail to do the same in human clinical trials [1]. This “translation failure” is one of the biggest challenges in biomedical research today: it is estimated that around two-thirds to 95% of therapeutics found to be safe and effective during animal testing, fail when tested in humans [1, 2, 3, 4]. Translatability has been defined as “the ability to apply research discoveries from experimental models to applications that directly benefit humans” [5]. Reasons for low translatability are multifaceted. However, the pervasiveness of suboptimal study design, analysis, and reporting, potentially resulting in a lack of reproducibility (i.e., the “extent to which the results of a study agree with those of replication studies” [5]), has been flagged as a key concern [2, 6]. Animal studies often demonstrate deficiencies such as inaccurate or inconsistent data collection procedures, poor reporting of key variables, including the age and sex of animals used, and a lack of measures to reduce risks of bias, including the absence of randomization or blinding [7, 8, 9]. In terms of statistical methodology, frequent issues include small sample sizes, leading to low powered studies, inadequate control for confounding variables, and insufficient description of statistical methods or reporting of uncertainty measures [10, 11, 12, 13]. Publication bias, the phenomenon in which the decision to publish a study is based on the direction or strength of its findings, is also rampant. Animal studies reporting positive and statistically significant results are more likely to be published than those with negative or statistically non-significant findings, meaning that subsequent human studies may be based upon biased conclusions [14, 15]. Such issues are a detriment to both animals, whose lives are wasted when incorrect conclusions are drawn from research performed on them; and humans, who are put at unnecessary risk during clinical trials when an intervention’s reported safety or efficacy is overstated or outright false [7, 16].

### Replication and translation

Recently, there has been a growing interest in replications of previously published studies. A replication is defined as a “study that repeats all or part of another study and allows researchers to compare their findings” [5]. To perform a replication study, researchers could for example use the same methodology and/or analysis as presented in an original study on newly collected data. They then attempt to determine if the results from the replication study are consistent with those in the original study [17]. A multitude of metrics have been used or proposed to quantify the consistency of results, and ultimately to decide if a replication was “successful” or not [18]. We will refer to these metrics as “replication success metrics”. They might compare, for example, the *p*-values or the magnitude, direction, or uncertainty of estimated treatment effects obtained from the original and replication studies. Other metrics, such as the one based on a meta-analysis, combine results from an original study and its replication attempt(s) to estimate an overall effect size [19, 20, 21]. So far, studies attempting to estimate how often translation failure occurs have largely utilized the simple statistical significance criterion, i.e., assessing if the animal and human studies both report a statistically significant treatment effect in the same direction, often referred to as the two-trials rule [22]. To our knowledge, the usage of alternative replication success metrics in a translation setting has not yet been investigated. Translation contrasts with replication in that animal and human studies examine different populations and often have different experimental designs, and thus inherently produce different results. As such, a human study is a “conceptual” rather than a “direct” replication of the animal study [23]. As a result, metrics which are useful for measuring replication may not be as applicable for translation. This distinction motivates the current simulation study.

In this paper, we define “translation success” statistically: a metric flags translation success when a certain condition, depending on the metric, is fulfilled. This condition reflects the translation goal, which could be, for example, confirming that a beneficial effect exists in both animals and humans, or assessing the similarity of effect sizes across animals and humans. This statistical definition is narrower than biological translation, which concerns the underlying mechanisms linking animal and human physiology. Several frameworks have recently been proposed to structure the use of preclinical evidence in decisions about progression to human trials [24, 25]. Quantitative tools that assess the consistency between animal and human efficacy findings, including the metrics evaluated here, can provide an empirical basis for one component of such frameworks.

### Study objectives

To date, little is known about the most appropriate metrics to assess or quantify “translatability” or “translation success”. In this study, we aimed to investigate whether metrics proposed to quantify replication success can be applied, and are useful, in the context of the translation of results from animals to humans. We investigated the ability of these metrics to quantify translation success under various simulation conditions, for example in different scenarios of effect sizes and sample sizes, in order to gain a better understanding of their behaviour and characteristics.

## 2 Methods

### 2.1 Study design, data, and protocol

This is a simulation study. Synthetic data sets, representing animal studies and human studies, were generated according to various simulation conditions (see Sections 2.3, 2.4 and 2.5). The simulated animal and human findings were then used to evaluate the selected translation success metrics (presented in Section 2.7) using pre-specified performance measures (Section 2.8). A detailed protocol of the present simulation study, following the ADEMP (Aims, Data-generating mechanisms, Estimands and other targets, Methods, Performance measure) preregistration template [26], was preregistered on the Open Science Framework prior to running the simulation study [27].

### 2.2 Protocol amendments

While drafting this manuscript, we found a conceptual error in our data generating mechanism. Initially, we planned to incorporate the heterogeneity variance directly into the simulation of the individual observations for the animal, respectively the human, study. We now first use the heterogeneity variance to simulate an effect size for the animal, respectively the human, study, and then use this effect size and solely the sampling variation to simulate the individual observations for the animal, respectively the human, study. Finally, a coding error in the protocol (in the calculation of the pooled variance the “-2” was missing in the denominator, page 4 of the protocol) led to the wrong human group sample size showing up in the protocol text (103 instead of the correct 107).

### 2.3 Data generation

We assumed that the synthetic studies investigated the effect of a treatment (*e.g*., prenatal amino acid supplementation) on an outcome that is comparable in both animals and humans (*e.g*., maternal blood pressure) as in Terstappen et al. [28]. We simulated individual observations of this outcome measurement for the animals, respectively for the humans, in the treatment group and the control group. To generate the synthetic data, the true animal and human means in the treatment groups were set to *µ*_A_ and *µ*_H_. The mean in the animal and human control groups was always set to 0, so that *µ*_A_ and *µ*_H_ would correspond to the mean difference in effect sizes. The true effect size heterogeneity variances were set to 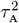 and 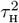. For each simulation repetition *i* we performed the following:

1. **Simulation of effect sizes:** We first simulated *k* animal effect sizes and one human effect size:

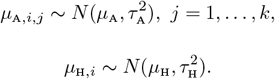
2. **Simulation of the animal finding:** We generated *k* synthetic animal studies. Each study *j* (*j* = 1, …, *k*) had *n*_A_ animals in the control group (C) and *n*_A_ animals in the treatment group (T). The outcomes were simulated as

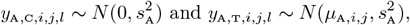

with *l* = 1, …, *n*_A_. For each of the *k* synthetic animal studies we then performed a one-sided two-sample *t*-test to compare the outcome for the treatment group to that of the control group. The *k* effect estimates were pooled using a random-effects meta-analysis (with restricted maximum likelihood estimator for heterogeneity variance), and the resulting pooled effect size, standard error and *p*-value constituted the “animal finding”.
3. **Simulation of the human finding:** We simulated outcome measurements for the human study, with *n*_H_ humans in the control group (C) and *n*_H_ humans in the treatment group (T):

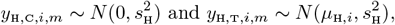

where *m* = 1, …, *n*_H_. A one-sided two-sample *t*-test was then performed on the simulated human outcome measurements to compare the treatment and control groups, and the resulting effect size (mean difference), standard error of the mean difference and *p*-value were extracted. This constituted the “human finding”.

We performed one-sided tests as we were specifically testing for a “beneficial treatment effect”, *i.e*., a decrease in maternal blood pressure. Note that we performed the one-sided tests with significance level *α* = 0.025, which is equivalent to performing two-sided tests at level 0.05, and checking that the effect goes in the beneficial direction.

### 2.4 Motivating dataset

The selection of the values of the simulation parameters in Section 2.3 was based on a systematic review and meta-analysis by Terstappen et al. [28] (see also Figure 2.1 in Huang and Heyard [27]), which assessed the effects of prenatal amino acid supplementation on birth weight and, as a secondary outcome, maternal blood pressure (BP, mmHg). In this simulation study, we focused only on the blood pressure data, for which measures from animal and human subjects were comparable. From this data we extracted the species-specific random-effects meta-analytical treatment effects, *θ*_A_ and *θ*_H_, and the estimated heterogeneity variances 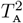 and 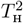. The typical within-study variances, 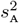 and 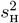, were also computed. The data included 15 animal studies and six human studies. The average sample size per group was 8.7 for the animal studies and 26.8 for the human studies. We acknowledge that simulation parameters based on a single meta-analysis may not be representative of the full breadth of biomedical research. Nevertheless, Terstappen et al. [28] was selected because it is one of a few meta-analyses to include both animal and human studies investigating the same research question. Further, the outcome (maternal blood pressure) chosen in this meta-analysis is comparable between animal and human studies, which is often not the case in the translation setting.

**Figure 1:**
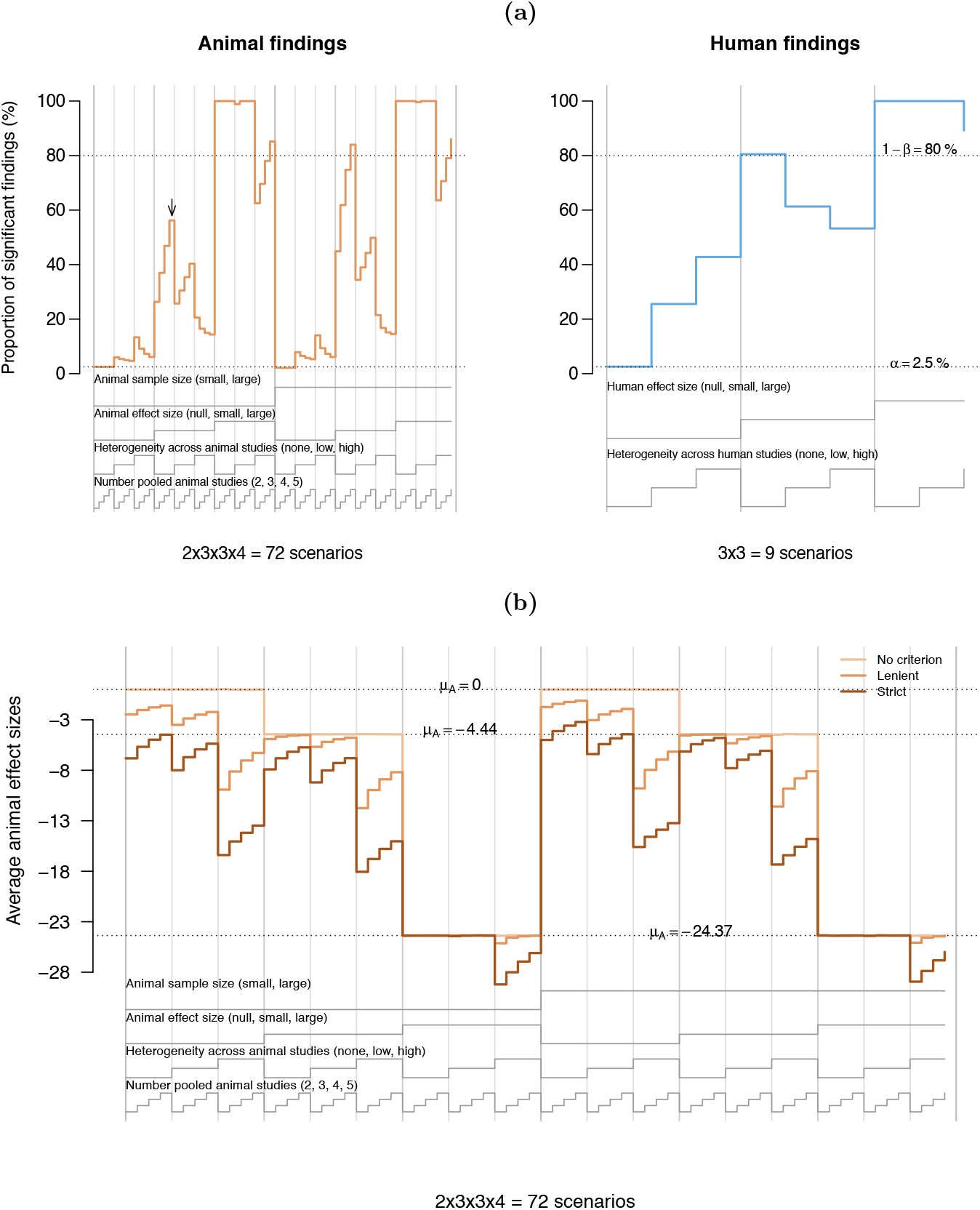
**(a)** Nested loop plot of the proportion of statistically significant animal and human findings over all simulation repetitions depending on the simulation conditions, i.e. animal and human effect sizes, heterogeneity across animal and human studies, animal study sample sizes, and the number of animal studies pooled together to obtain the animal finding. The dotted horizontal lines represent a proportion of 2.5% and 80%. The legend under each plot shows which of the progressively thinner columns in the plot correspond to which combination of simulation conditions. Each horizontal line segment contains the proportion of significant findings under each combination of conditions. For example, **the segment highlighted with the arrow** represents the proportion of significant findings in the animal studies, when the smaller sample size was used, the animal effect size was small, there was no heterogeneity across the animal studies, and 5 animal studies were pooled. **(b)** Nested loop plot of the average animal effect size over all simulation repetitions, depending on the simulation conditions and the decision criterion applied on the animal finding. The average effect size observed in the human studies is not affected by the applied criterion. Note that since the criterion was not added as a simulation condition, the represented data is correlated, as the same simulation repetitions are used to calculate the average effect size for the strict, lenient and no criterion.

**Figure 2:**
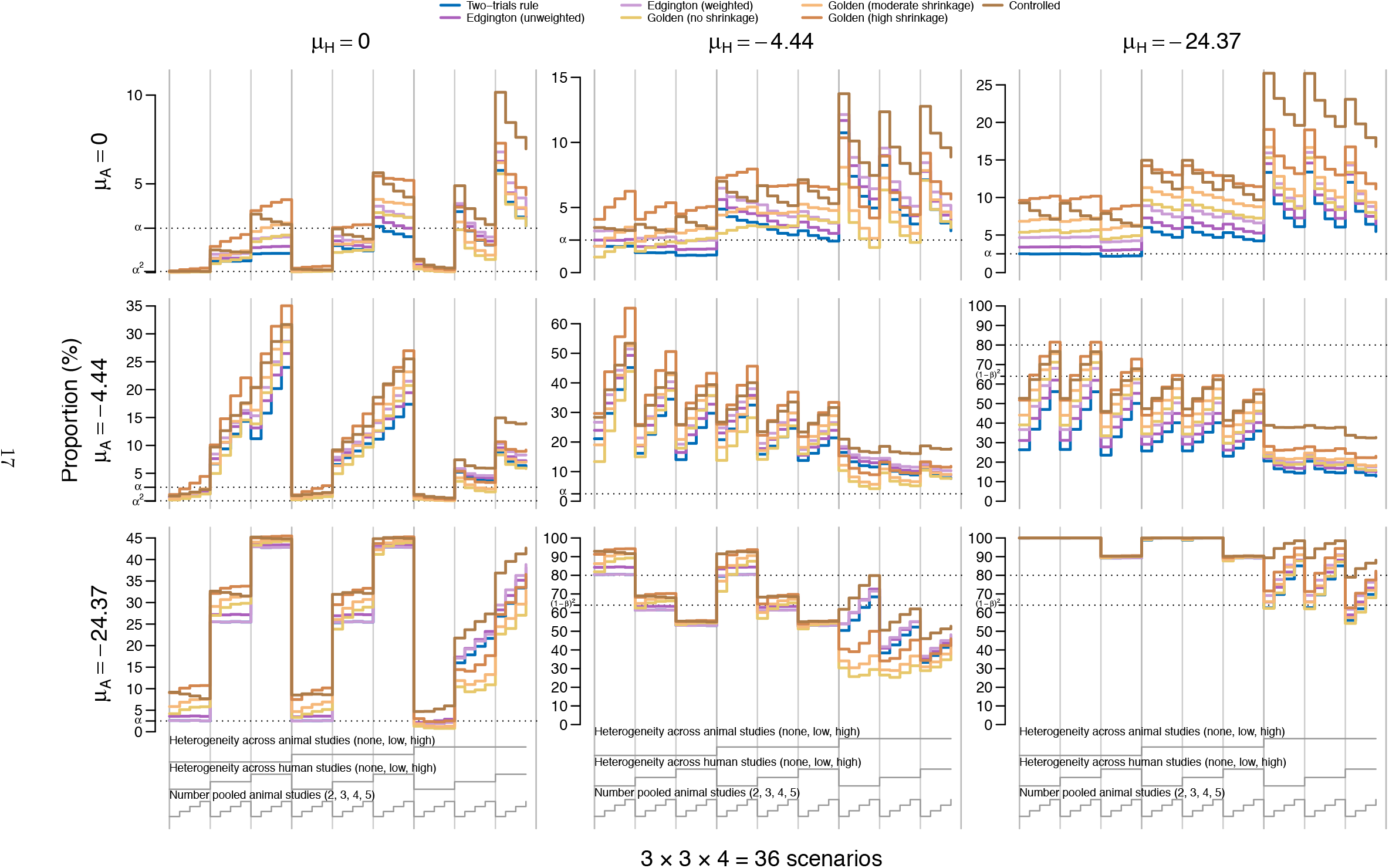
Grid of nested loop plots of the proportions of animal-human pairs for which the different metrics flagged successful translation across simulation conditions under no criterion. Each of the plots in the grid represent another animal-human finding combination. In the first column, for example, the human studies are all simulated under the null hypothesis of no effect. Note that the results for the replication BF and the meta-analysis are not shown here for better readability. The dotted horizontal lines represent *α*^2^ = 0.000625, *α* = 0.025, 1 − *β* = 0.8 and (1 − *β*)^2^ = 0.64. All animal studies in this representation are simulated with a small sample size per group (*n*_A_ = 10).

### 2.5 Simulation conditions

To investigate the applicability of the replication success metrics in the scenario of animal to human translation, we simulated the animal and human findings under various conditions described below. The conditions were chosen based on previous literature [7, 13] and expert knowledge, with the aim of emulating plausible real-world translation scenarios as closely as possible.

#### Animal and human effect sizes, *µ*_A_ and *µ*_H_

In the motivating dataset, we found that *θ*_A_ = −24.37 mmHg and *θ*_H_ = −4.44 mmHg. This means that a beneficial treatment effect (i.e., a reduction in blood pressure) was found in both animals and humans, but the effect was larger in animals. We additionally simulated scenarios in which the true treatment effects in animals and humans were the same, *i.e*., both small, both large, or both absent entirely. Alternatively, although less plausible in practice, the true effect size in humans could be larger than in animals. We therefore simulated under all possible combinations of small, large, or zero animal and human effect sizes, summarized in Table 1. These effect sizes span a clinically meaningful range on the unstandardised mean difference scale for blood pressure measurements: from no effect to a modest but clinically relevant reduction (−4.44 mmHg) to a large effect which is more typical for animal studies (−24.37 mmHg^1^). Note that we did not investigate cases where the true animal and human effects go in opposite directions, *i.e*., where the treatment is beneficial for one species but harmful for the other.

**Table 1:**
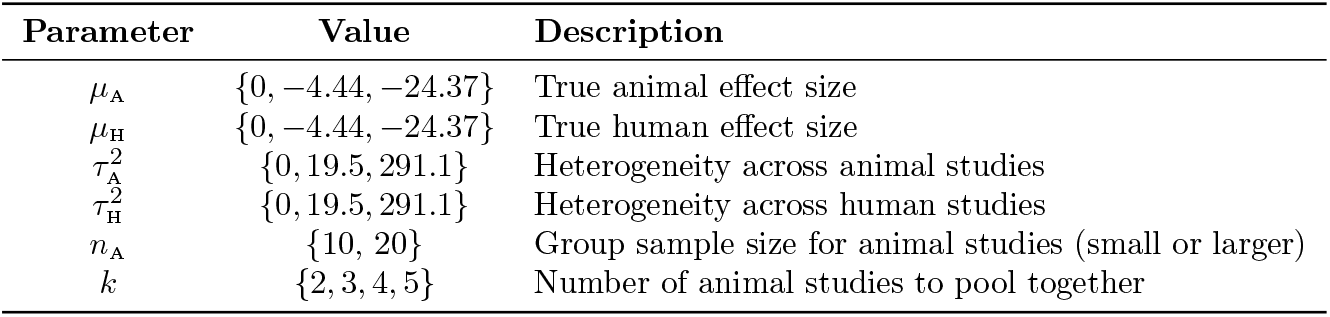
Summary of the simulation factors used to generate animal and human studies. These were varied in a fully factorial manner.

| Parameter | Value | Description |
| --- | --- | --- |
| $\mu_A$ | $\{0, -4.44, -24.37\}$ | True animal effect size |
| $\mu_H$ | $\{0, -4.44, -24.37\}$ | True human effect size |
| $\tau_A^2$ | $\{0, 19.5, 291.1\}$ | Heterogeneity across animal studies |
| $\tau_H^2$ | $\{0, 19.5, 291.1\}$ | Heterogeneity across human studies |
| $n_A$ | $\{10, 20\}$ | Group sample size for animal studies (small or larger) |
| $k$ | $\{2, 3, 4, 5\}$ | Number of animal studies to pool together |

#### Animal and human study heterogeneity, 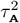 and 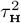

We implemented a similar setup for the between-study heterogeneity variances. In our motivating dataset, animal studies had a higher degree of heterogeneity 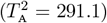 than human studies 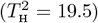. The large heterogeneity value corresponds to a relative heterogeneity of *I*^2^ of 95.33% in the animal meta-analysis. While this might appear high, Hooijmans et al. [30] showed that 55% of animal study meta-analyses using raw mean differences as effect size measure have *I*^2^ *>* 75%, many also have *I*^2^ closer to 100%. Similarly to the simulation of effect sizes, we simulated under all possible combinations of small (i.e., 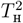), large (i.e., 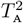), or zero animal and human study heterogeneity (see Table 1). Again, our simulation conditions span a meaningful range of possible levels of heterogeneity: from none to moderate to more extreme levels.

#### Animal and human study sample sizes, *n*_A_ and *n*_H_

Animal preclinical studies commonly suffer from insufficient sample sizes, leading to underpowered studies [2, 14, 6]. Therefore, we included two different sample sizes per group in each of the simulated animal studies. The typical sample size observed in animal studies is small, with approximately 10 animals per group [28, 10, 31]. To investigate the effect of an increased animal sample size on translation success, we simulated with a larger sample size of 20 animals per group. This represents the maximum number of animals per group observed in our data example from Terstappen et al. [28] and in Table 1 of Hooijmans et al. [31]. For the simulated human studies, we used a fixed sample size of *n*_H_ = 107 humans per group. This sample size was chosen to achieve 80% power for an absolute effect of |*θ*_H_| = 4.44, typical within-study variance 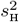, and a significance level *α* = 0.025, using a one-sided two-sample *t*-test.

#### Number of pooled animal studies, *k*

In real-world drug development, multiple animal studies are usually performed and results are pooled before deciding to progress to a human study. We investigated the effect of pooling together different numbers of animal studies on translation success: *k* = 2, 3, 4, and 5. While in practice, more than five animal studies might be conducted and pooled, we expect that varying *k* from 2 to 5 will give us enough of an indication of how increasing *k* affects translation and the performance of the translation success metrics. As described above, we performed a random-effects meta-analysis (using the restricted maximum likelihood estimator for the heterogeneity variance) of the *k* animal studies to generate the animal finding.

We varied the factors above, summarized in Table 1, in a fully factorial manner. This resulted in 3 (animal effect size) × 3 (human effect size) × 3 (animal heterogeneity) × 3 (human heterogeneity) × 2 (animal sample size) × 4 (number of animal studies to pool together, *k*) = 648 simulation conditions in total.

### 2.6 Criteria to continue from animal to human studies

Usually, animal studies must show evidence of a beneficial treatment effect before a treatment is tested in humans. Alternatively, treatments that show no evidence for a beneficial effect in animal studies may continue to testing in humans if the treatment is safe and its mechanism of action is plausible in humans [32, 33]. When analysing the applicability of translation success metrics, we considered both of these continuation criteria and excluded the human studies accordingly. Table 2 shows the definition of the three decision criteria implemented in our simulation, together with a description of what their real-world analogue could be.

**Table 2:** Definition and rational for the implemented decision criteria to continue.

| Name | Description | Real-world analogue |
| --- | --- | --- |
| **Strict criterion** | We progressed to a human study if the random-effects meta-analysis of the corresponding $k$ animal studies found a significant beneficial treatment effect with one-sided significance level $\alpha = 0.025$. | Related to regulatory-style evidence thresholds and presents a highly selective preclinical pipeline. |
| **Lenient criterion** | We progressed to a human study whenever the random-effects meta-analysis of the $k$ animal studies found a beneficial effect (*i.e.*, if the estimated effect was negative), even if it was not statistically significant at $\alpha = 0.025$. | Represents an exploratory decision-making where efficacy evidence is considered in addition to other factors (*e.g.*, safety), but not intended to represent a certain regulatory standard. |
| **No criterion** | As a point of reference, we also computed performance measures for all simulated animal and human findings, regardless of the results of the animal studies. | Represents a reference scenario mainly used to evaluate metric behaviour independent of progression decisions. |

### 2.7 Translation success metrics

We compared the characteristics of nine translation success metrics, including the replication success metrics used in Freuli, Held, and Heyard [19] and Muradchanian et al. [20] as well as some more recently developed metrics, across the previously defined simulation conditions: the significance criterion, the meta-analysis, the replication Bayes factor, the unweighted and the weighted Edgington’s method, the controlled sceptical *p*-value and three versions of the golden sceptical *p*-value.

These metrics were primarily designed to assess replication success in the pairwise comparison of one original study with one replication study, where both studies were conducted using the same methodology in the same population. Translation differs in that the same research question is investigated across different populations. To investigate the applicability of replication success metrics to the translation setting, the pooled result of the random-effects meta-analysis of *k* animal studies was treated as “original finding” (*i.e*., the animal finding), while the result of the corresponding human study was treated as the “replication finding” (*i.e*., the human finding).

#### Significance criterion (Two-trials rule)

The significance criterion, often referred to as the two-trials rule, is the current standard for a new drug to meet prior to its approval. It requires that two independent studies demonstrate a drug’s efficacy at a certain significance level, usually *α* = 0.025 for one-sided hypothesis testing [22] to take into account direction of effect. This criterion is also often used to identify replication success in large-scale reproducibility projects [18]. According to the significance criterion, we flagged a successful translation if both the animal and human studies yielded evidence for a beneficial treatment effect, both at a significance level of *α* = 0.025 [20]:

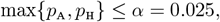

where *p*_A_ and *p*_H_ represent the *p*-values found in the animal and human studies, respectively. By setting *α* = 0.025, the significance criterion controls the overall T1E rate, or in our case the rate of a false positive translation success, at *α*^2^ = 0.025^2^ = 0.000625 [22]. Note that this metric treats the animal and human finding as interchangeable.

#### Meta-analysis

According to the meta-analysis criterion, we flagged translation success if a fixed-effects meta-analysis combining the animal and the human findings found a significant effect in the desired direction (here, a decrease), at a one-sided significance level *α*^2^, i.e., *p*_MA_ *< α*^2^. This threshold again ensured an overall T1E control at *α*^2^ [34]. We followed Freuli, Held, and Heyard [19] for the implementation of the method via the weighted version of Stouffer’s method described in Cousins [35], and define the meta-analytic *p*-value *p*_MA_ of a one-sided test for a negative effect (i.e., a beneficial effect) as follows:

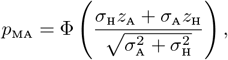

where Φ(·) is the standard normal cumulative distribution function, and *z*_A_ and *z*_H_ are the *z*-values representing the findings in the synthetic animal studies (pooled) and human study. If we were to test for a positive effect, the formula for the desired *p*-value would change to 1 − Φ(…). Note that we used fixed-effects meta-analysis as this represents the commonly used metric in the replication context. Alternatively, we could have used random-effects meta-analysis, but the assessment of heterogeneity is challenging when only two findings (animal and human) are considered [36]. The metric based on meta-analysis treats the animal and human finding as interchangeable.

#### Replication Bayes factor

In the translation setting, the replication Bayes factor (BF) quantifies the evidence that the outcome observed in a human study is absent or spurious (*H*_0_) relative to the evidence that the outcome in the human study is consistent with that found in the (original) animal studies (*H*_*r*_) [37]. To calculate the replication BF, BF_0r_, *H*_*r*_ is defined as the alternative hypothesis that the human effect is distributed according to the posterior distribution of the effect after observing the animal finding. A translation was flagged as successful if

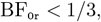

using the conventional threshold for substantial evidence for *H*_*r*_ over *H*_0_ [38].

#### Unweighted & weighted Edgington’s method

Edgington [39] developed an additive method of combining *p*-values from independent experiments, which has been applied more recently in a replication success setting [40]. Under the original version of Edgington’s method, to control the overall T1E rate across two studies at level *α*^2^ = 0.025^2^, a successful replication is flagged if the sum of the *p*-values in the original study *p*_*o*_ and the replication study *p*_*r*_ is smaller than 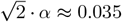. With Edgington’s method, it is possible to flag success even if one of *p*_*o*_ or *p*_*r*_ is not significant, as long as *p*_*o*_ + *p*_*r*_ ≤ 0.035. In our study, a successful translation was flagged with Edgington’s method if

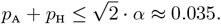

Even more recently, a weighted version of Edgington’s method has been proposed [40]. Here, an original study is down-weighted and a replication study is up-weighted to account for potential biases in the original study, and study findings are no longer interchangeable. For the same overall T1E control at level *α*^2^ = 0.025^2^, and in the case in which a replication study carries twice the weight of the original study, a successful replication is flagged if *p*_*o*_ + 2*p*_*r*_ ≤ 2 · *α* = 0.05 [40]. In our study, we gave the human result more weight than the animal finding, and flagged a successful translation with weighted Edgington’s method if

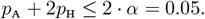

#### Golden & controlled sceptical *p*-value

The sceptical *p*-value combines a reverse-Bayes technique with a prior-data conflict assessment. The extent to which the data in a replication or human study conflicts with a sceptical prior which renders the original or animal finding not convincing, can be quantified with the sceptical *p*-value [41, 20]. In our study, we will examine two specific versions of the sceptical *p*-value: the golden and controlled sceptical *p*-value [42, 43].

With the golden sceptical *p*-value *p*_*s*_, success can be flagged if the *p*-value of the animal finding is sufficiently small (i.e., *p*_A_ ≈ *α*), even if it is not necessarily significant, as long as there is no shrinkage in effect size in the human study [42, 19]. However, in the translation setting, shrinkage of effect size is expected in human studies relative to animal studies. Held, Micheloud, and Pawel [42] have developed a method to calculate a threshold 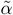 for the golden sceptical *p*-value in the presence of shrinkage, which is equivalent to a pre-specified threshold of *α* when no shrinkage is present. The golden sceptical *p*-value controls the overall T1E rate at a maximum level of *α*^2^, provided that the sample size of the replication or human study is larger than in the original or animal study. We can therefore calculate the values for 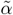 below which a successful translation can be flagged even if *p*_A_ is sightly higher then *α*, in the presence of no (0%) shrinkage, moderate (25%) shrinkage, and high (50%) shrinkage. Note that these levels of shrinkage were selected rather arbitrarily and different levels of shrinkage could be selected as needed. We based them on an observed shrinkage of about 50% in the Replication Project Psychology [44], while it was even higher in the Replication Project Cancer Biology (mean effect size of 6.15 in the original studies and 1.37 in the replication studies) [45]. In our study, translation success was flagged if the following conditions were satisfied, depending on the allowed level of shrinkage:

1. 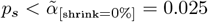 when allowing for no shrinkage;
2. 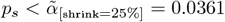 when allowing for moderate shrinkage; or,
3. 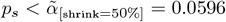 when allowing for high shrinkage.

Finally, the controlled sceptical *p*-value 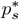, another recalibration presented in Micheloud, Balabdaoui, and Held [43], guarantees control of the overall T1E rate at level *α*^2^. Translation success was flagged if

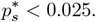

Note that whenever the animal finding indicated a harmful effect (i.e., the effect goes in the opposite direction than what was expected), we implemented all sceptical *p*-value metrics in a way that forced them to flag failure. This approach is valid, as no version of the sceptical *p*-value would ever flag success if the original or the animal finding has a very high *p*-value.

### 2.8 Performance measures

To evaluate the performance of each translation success metric, we calculated and compared the proportion P of synthetic pairs of animal and human findings for which the metric flagged successful translation under the different simulation conditions. The denominator for this proportion depends on the animal finding (i.e., the results of the meta-analysis of *k* animal studies) and the different continuation criteria (strict, lenient, none) described in Section 2.6. This leads to the following three versions of P (where N stands for number):

- Under the strict criterion: 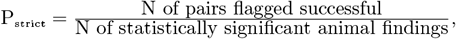,
- Under the lenient criterion: 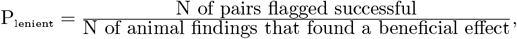,
- Under no criterion: 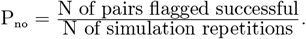.

Under the assumption of animal and human null effects, this proportion reflects the overall T1E rate – the rate of *false positive* translation success. The lower this proportion, the better a metric is at uncovering false translation failure. Under the assumption that both animal and human effects are not null, the proportion can be interpreted as translation power, i.e., the probability of *true positive* translation success. The higher the proportion of true positive translation success, the better the metric is suited to “correctly” declare translation success under the chosen simulation conditions.

We used so-called “nested loop plots” to represent and compare the proportions for the different metrics across simulation conditions as recommended by Rücker and Schwarzer [46]. The combinations of simulation conditions are ordered and arranged on the horizontal axis, while the proportion of successful translations is presented on the vertical axis^2^.

### 2.9 Monte Carlo uncertainty and number of simulation repetitions

The number of simulation repetitions was calculated based on a maximum desired Monte Carlo standard error (MCSE) of 0.5% for P [47]. We considered the “worst-case” scenario of P = 0.5 (i.e., the metric is not better than tossing a coin), as well as the strictest criterion for a human study to be performed.

From this, we obtained a maximum of 400^′^000 animal studies to simulate in order to move on to at least 10^′^000 human studies, while maintaining a maximum MCSE of 0.5%. For simplicity, we simulated 400^′^000 animal studies under all combinations of simulation conditions.

### 2.10 Implementation

Our simulation study was implemented in `R` (version 4.5) and designed using the `SimDesign` package [48]. We used the `BFr` function from the `BayesRep` package to compute the replication BF [49], and the `ReplicationSuccess` package for all versions of the sceptical *p*-value [50]. Following Pawel et al. [51], we recorded and reported the proportion of missingness. This is a common issue in simulation studies in which problems such as non-convergence of optimization algorithms may cause some simulation repetitions and conditions to yield invalid outputs, leading to missing values for the performance measures.

## 3 Results

### 3.1 Characteristics of simulated animal and human studies

We first illustrate the impact of our simulation design choices on the animal and human studies separately, and verify that the simulations were performed as expected. For this, Figure 1.(a) shows a nested loop plot with the proportion of significant animal and human findings (one-sided *p < α* = 0.025) according to the simulation conditions.

As expected, both animal and human studies show a T1E rate (i.e., proportion under the null) of about *α* = 0.025 under the null hypothesis of no effect (i.e., *µ*_A_ = 0 and *µ*_H_ = 0) combined with no heterogeneity across studies. Increasing heterogeneity increases the T1E rates. For the animal findings, the T1E rate decreases with an increasing number of pooled studies *k*.

The human study sample size of *n*_H_ = 107 was chosen in order to achieve 80% power assuming a small effect size and no heterogeneity. Accordingly, under these conditions, we also find that about 80% of the human findings are significant. Also as expected, the power decreases with increasing heterogeneity. Simulating under the large human effect using the same sample size *n*_H_, naturally results in power close to 1, except when heterogeneity is large.

On the other hand, the animal findings have low power when under the simulation condition of a small animal effect. As expected, power increases with increasing *k* and the larger animal sample size, but still remains rather low. Increasing heterogeneity across the animal studies further lowers the proportion of significant animal findings. Finally, simulating under the large animal effect results in highly powered findings, almost 100% power, except in the case of high heterogeneity.

Then, Figure 1.(b) shows that conditioning the decision to conduct a human study on the animal finding (being beneficial or significant) results in overestimated effect sizes for the animal finding. The stricter the decision criterion, the more inflated the estimated average effect size in the animal studies.

#### Missingness

Our simulation study was also affected by missingness, though only in rare cases. Specifically, missingness occurred only in the data generating mechanism (see classification in Pawel et al. [51]) and was due to non-convergence of the Fisher scoring algorithm of the meta-analysis of the animal studies using the `rma` function. A table in our online appendix (https://rachelheyard.pages.uzh.ch/translation_simulation/) summarizes the proportion of missingness for each combination of simulation condition, with a maximum of 0.0025% (i.e., 10 missing values out of 400’000 repetitions). These repetitions were omitted in analyses.

### 3.2 Performance of translation success metrics

Figure 2 shows the proportion of animal-human pairs for which the different metrics flagged successful translation across simulation conditions. The figure specifically shows the proportion P_no_ (no criterion) with small animal group sample size, *n*_A_ = 10. Note that Figure 2 does not include the results for the replication BF and the meta-analysis for readability reasons as they behave very differently (see the corresponding Figure A.1 in the appendix). Our online appendix allows the reader to zoom into the different plots and also contains the results with the larger animal sample size; see https://rachelheyard.pages.uzh.ch/translation_simulation/. A less technical summary of the main results listed below can be found in Table 3.

**Table 3:** Summary of simulation results across 9 translation success metrics (no criterion). *At baseline* refers to no heterogeneity across animal and none across human studies. The proportion P refers to the proportion of pairs of animal and human findings for which the metric flagged translation success (under specific simulation conditions).

| Metric | Strengths | Weaknesses | Behaviour with increasing heterogeneity | Sensitivity to more animal data (larger $k$ and $n_A$) | Behaviour under effect mismatch (animal vs human) |
| --- | --- | --- | --- | --- | --- |
| **Significance criterion** | Excellent control of overall T1E rate at $\alpha^2$ (at baseline); one of the most reliable metrics under heterogeneity in human studies. | Lowest translation power in many scenarios; often too conservative when animal findings are low-powered. | Tends to be the metric least affected compared to baseline, unless human heterogeneity is high. | Larger $n_A$ increases P slightly; larger $k$ decrease false translation success rate and increase translation power, unless there is high animal heterogeneity. | Independent; it requires animal and human study to be significant; a large animal effects cannot "mask" a null human effect. |
| **Meta-analysis** | Highest translation power in many conditions; outperforms other metrics when true effects exist in both species. | Very high overall T1E rates (up to 40%); flags success even if only one species shows a strong result. | Highly sensitive to human heterogeneity (high heterogeneity in human causes a massive increase in false positive translations); less sensitive to animal heterogeneity. | Larger $n_A$ and $k$ increase P if animal effect is non null, and decrease it otherwise. | Treats animal and human as interchangeable and findings as equivalent, allowing a very convincing animal finding to hide a human null result (and vice-versa). |
| **Replication BF** | Useful in case of high heterogeneity scenarios (where it becomes more comparable to other metrics). | Very low power under no to low heterogeneity scenarios; potentially weights human data too heavily. | Very sensitive to changes in heterogeneity; P generally increases with heterogeneity, unless animal effect is null or small and human is small. | Larger $n_A$ decreases P; an increase in $k$ increases P when true animal and human effect are the same, and decreases it if there is a mismatch, unless there is high heterogeneity in animal study (which inverts the trend). | Human-focused; functions a bit like a test of the human study, largely ignoring the original animal evidence. |
| **Edgington** (*unweighted / weighted*) | Better translation power than significance criterion while maintaining good overall T1E rate control at baseline; P for weighted version pulled up or down depending on evidence in human studies. | Generally conservative; weighted version can have slightly higher overall T1E rates than significance criterion. | Generally follows significance criterion with slightly higher proportions; unweighted and weighted come closer together with high animal heterogeneity. | Larger $n_A$ generally increases proportions unless there is no heterogeneity and a null effect in the animal studies; proportions increase slightly with $k$ if animal effect is non zero. | Additive; requires evidence from both but allows a very strong result in one to compensate for a slightly weaker result in the other. |
| **Golden sceptical p-value** (*No shrinkage*) | Best at keeping overall T1E rates low when animal study heterogeneity is high. | Lowest translation power among sceptical $p$ metrics and lowest translation power in small effect scenarios; penalizes any discrepancy in effect size. | Sensitive, especially to high heterogeneity across animal studies. | Translation power increases with $k$ due to higher chance of a significant animal finding unless there is high animal heterogeneity; P increase with $n_A$. | Penalizing; specifically designed to flag failure if the human effect is smaller than the animal effect (shrinkage) and one of the studies is not convincing enough. |
| **Golden sceptical p-value** (*Mod/High shrink*) | High translation power; allows weaker animal findings to translate when human findings are strong and vice-versa, even if shrinkage is present. | Higher overall T1E rates; less conservative to shrinkage (by design). | Sensitive to human heterogeneity; similar trend compared to no shrinkage version. | Highly sensitive; translation power increases significantly with $k$ as it decreases the relative sample size ratio; P increase with $n_A$. | Adaptive; permits for potentially realistic shrinkage between animal and human effects without automatically flagging failure. |
| **Controlled sceptical p-value** | Most consistently high translation power across varying conditions. | Higher overall T1E rate than significance criterion; similar overall T1E patterns to high-shrinkage metrics. | Moderate sensitivity; follows the overall T1E rate of human studies as heterogeneity increases. | Strong response to $k$; increasing $k$ to 5 allows the metric to reach acceptable translation power even with noisy animal data; P increase with $n_A$. | Balanced; allows for effect size differences but requires a convincing level of evidence that accounts for the sample size ratio. |

#### Assuming a large animal effect and a small human effect

(bottom center plot in Figure 2) This combination of animal and human effect sizes is closest to the results from the meta-analysis in Terstappen et al. [28], and is therefore potentially the most realistic in the translation setting. Here, a well-performing metric should find a relatively high proportion of translation successes, i.e., high translation power.

When no heterogeneity is present in either animals or humans, the translation power for all metrics in the figure is at least 1 − *β* = 80%. The two-trials rule and the weighted Edgington are both equal to 80% (lines overlap). Unweighted Edgington behaves similarly with a sightly higher proportion. The controlled and golden (high shrinkage) sceptical *p*-values find the highest translation power.

Increasing the heterogeneity in human studies decreases the translation power of all metrics and brings them closer together. When heterogeneity of the human studies is low and animal heterogeneity is none, all metrics are close to (1 − *β*)^2^. These results barely change when increasing the animal study heterogeneity from none to low. This might be due to the fact that the relative sample size *c* = *n*_H_*/n*_A_ is always larger than 1, even if the sample size of the animal finding is artificially increased with increasing *k*. A *c >* 1 forces some metrics to give more weight to the human study; therefore, even slight increases in the heterogeneity across human studies affects translation results. The translation power of the three golden sceptical *p*-values generally increases with *k*, as a higher *k* leads to a higher chance of observing a significant animal finding. The translation power of the three golden sceptical *p*-values is lowest when animal study heterogeneity is high, which can be explained by the decrease in power of the individual animal studies. The controlled sceptical *p*-value has a similar pattern with respect to *k* and animal heterogeneity, but increasing *k* to 5 countermeasures and its translation power is equal to 80%.

The metric based on a meta-analysis outperforms all other metrics in most conditions represented in Figure A.1. From previous research [19, 43], we know that if either the animal or the human finding is convincing, the likelihood for the meta-analysis to flag success is high, regardless of the evidence in the other study. The replication BF results in very low proportions of successful translation when there is no or low heterogeneity across animal and human studies, because the effect sizes from animal and human findings are too inconsistent. The results for the replication BF are more comparable to the results of the other metrics in the presence of high study heterogeneity in either animals or humans.

Under the lenient criterion, the conclusions are the same. Under the strict criterion, most conclusions hold, while the proportions for the two-trials rule, both Edgington and the controlled *p*-value are now independent from increases in *k* and increases in the level of heterogeneity across animal studies. Larger animal sample size per group increases all proportions slightly, apart from the proportion for the replication BF, which decreases.

#### Assuming null effects in animal and human studies

(upper left corner in Figure 2) Here, a well-performing metric should find a low proportion of translation successes, i.e., a low overall T1E rate or false positive translation success rate. All metrics except the replication BF in (Figure A.1) control the overall T1E rate at *α*^2^ when there is no animal or human study heterogeneity. An increase in human study heterogeneity inflates the proportion of false positive translations. The two-trials rule is least affected by changes in heterogeneity across human studies, followed by the unweighted Edgington, weighted Edgington and controlled sceptical *p*-value. The golden sceptical *p*-value (no shrinkage) keeps the overall T1E rate low, especially when there is no heterogeneity across studies of either animals or humans, which aligns with the theoretically expected pattern [42]. The golden sceptical *p*-values allowing for moderate/high shrinkage permit weaker animal findings to translate even if shrinkage is observed in the human study, but raise the overall T1E rate.

More animal study heterogeneity also increases the proportion of translation successes for all metrics. For all golden sceptical *p*-values, the increase in the proportion with *k* when there is no animal study heterogeneity is due to the corresponding decrease in relative sample size *c*. However, when there is low or high animal study heterogeneity, an increase in *k* leads to a decrease in the overall T1E rate for all metrics in Figure 2. This is likely related to the fact that increases in *k* decrease the partial animal T1E rate when animal study heterogeneity is low or high (see Figure 1.(a)). Generally, when there is no or low heterogeneity across animal studies combined with any level of heterogeneity across human studies, the two-trials rule performs relatively well, with the overall T1E rate mostly below *α*, and the weighted Edgington follows closely behind. However, when the heterogeneity across animal studies is high, the golden sceptical *p*-value (no and moderate shrinkage) performs better compared to the other metrics.

The metric based on meta-analysis and the replication BF, visible in Figure A.1, results in very high overall T1E rates. The replication BF weights the evidence of the “replication”, i.e., the human study, more heavily than the “original” animal finding. Increases in human study heterogeneity increase the partial T1E rate of the human finding; likewise, the same is true for the overall T1E rate in the case of the replication BF. The meta-analysis metric treats the animal and human findings as interchangeable and it is therefore enough if just one of the findings is very convincing to flag success. Since high heterogeneity in human studies results in an increased risk of a false positive human result, the overall T1E rate of the meta-analysis metric increases as well, up to 40% in extreme cases. Neither the replication BF nor the meta-analysis metric are affected much by increases in animal study heterogeneity. An increase in *k* decreases the overall T1E for the meta-analysis slightly, while it increases the overall T1E for the replication BF, especially when there is high heterogeneity across animal studies.

Results under the lenient criterion can be studied in the online appendix and follow similar trends. Under the strict criterion, translation success is conditional on the animal finding being significant. Consequently, the two-trials rule, both versions of Edgington’s method and the controlled sceptical *p*-values, which previously controlled the overall T1E rate at *α*^2^ when there was no study heterogeneity in either animals or humans, now do so at level *α* = 0.025. The golden sceptical *p*-value (no shrinkage) now yields the lowest translation success rates across all scenarios. The results for the replication BF and the meta-analysis are more comparable to those of the other metrics, aside from the replication BF when human study heterogeneity is high. Under the strict criterion, the overall T1E tends to increase with *k*, except when there is high heterogeneity across animal studies.

#### Assuming small animal and human effects

(center plot in Figure 2) Here, we observe translation success rates that are much lower than what we would expect, i.e., *<* (1 − *β*)^2^ = 0.64, except for the replication BF in Figure 1.A. This is due to the fact that the animal studies with *n*_A_ = 10 have insufficient power to detect a small effect. As shown in Figure 3.A, the results look slightly better with the larger animal study sample size. In addition, by artificially increasing the sample size of the animal finding with *k*, we also observe that the rates for all metrics increase at least slightly. Notably, however, this trend is reversed when there is high heterogeneity across animal studies. Increases in heterogeneity across studies of any species decreases the translation power for all metrics except the replication BF and meta-analysis.

The replication BF results in the highest proportion of successful translations under all conditions, generally followed by the meta-analysis. This can be explained by the fact that the replication BF puts more weight on the human study, and meta-analysis treats human and animal findings as interchangeable. Among the golden sceptical *p*-values, the version allowing for no shrinkage would be the most appropriate here, since the true animal and human effect sizes are assumed to be equal. This metric leads to the lowest translation power, apart from when the heterogeneity across human studies is high; then, the two-trials rule leads to similar or smaller translation success rates. The controlled sceptical *p*-value generally yields higher translation power compared to the other metrics, and is even the highest when there is high animal study heterogeneity, aside from the replication BF and meta-analysis. The golden sceptical *p*-value (high shrinkage) also performs similarly well, especially with no or low animal study heterogeneity.

Under the strict criterion (see online appendix), the two-trials rule, unweighted and weighted Edgington and the controlled sceptical *p*-value are approximately equivalent to the power of the human studies (1 − *β* = 0.8), as illustrated in Figure 1.(a). The golden sceptical *p*-value (no shrinkage) with borderline significant results generally finds the lowest proportion of translation success across conditions. The low-powered animal studies might lead to overestimated effect sizes for the animal findings, which is most penalised by this version of the golden sceptical *p*-value (no shrinkage). In addition, weighted Edgington finds proportions of translation success that are equivalent or slightly smaller than those of unweighted Edgington, while the proportions are larger when applying no criterion. This is because weighted Edgington puts less weight on the animal findings, which are heavily inflated under the strict criterion.

#### Assuming large animal and human effects

(bottom right plot in Figure 2) When applying no criterion, most metrics find a translation power of almost 100% under most simulation conditions, except when there is high heterogeneity across animal studies, as in that case the animal studies have low power. Translation success rates are even closer to 100% under the strict criterion.

#### Assuming a small animal effect and a human null effect

(center left plot in Figure 2) Under this combination of effect sizes, translation success should not occur. Indeed, the translation success rates for all metrics are generally small (*< α*) when there is no heterogeneity across animal or human studies. However, the proportion increases substantially with increasing levels of heterogeneity across human studies, and decreases with increasing levels of heterogeneity across animal studies. The meta-analysis, replication BF, sceptical *p*-value (high shrinkage) and controlled sceptical *p*-value lead to the highest proportions, while the golden sceptical *p*-value (no shrinkage) and the two-trials rule lead to the lowest proportions. Note that the animal studies have low power to detect a small effect.

#### Assuming an animal null effect and a small human effect

(top center plot in Figure 2) When the human effect is small, for which the human studies were powered at 80%, and the animal effect is null, all metrics but the replication BF and the meta-analysis generally result in translation success rates close to *α*. The proportion for the replication BF is close to the power of the human studies. Under the strict criterion, all metrics get closer to the human study power. Interestingly, under the strict criterion, the metric based on the meta-analysis is one of the metrics with the lowest proportions. This might be due to the human studies being powered at 80% and not higher.

#### Assuming a large animal effect and a human null effect

(bottom left plot in Figure 2) Here, the animal studies have high power, though power decreases with increasing heterogeneity across animal studies. Accordingly, when animal study heterogeneity is non-existent or low, there is a high chance of a very convincing animal finding, which then results in high translation success rates for the meta-analysis metric. The replication BF tends to be the most conservative unless there is high animal study heterogeneity. The remaining metrics all perform similarly well, are only slightly affected by changes in *k* and generally follow the partial T1E rate of the human studies.

#### Assuming a small animal effect and a large human effect

(center right plot in Figure 2) Here, the human studies are highly powered and the animal findings have low power. Hence, applying the strict criterion (see online appendix) leads to a translation success rate of almost 100% for all conditions except when heterogeneity across human studies is high, where it is then close to 90%. When no criterion is applied, the high-powered human studies still lead to a proportion of 1 or close to 1 for the replication BF and the meta-analysis. The remaining metrics are more conservative, with the two-trials rule yielding the smallest proportions across conditions. Proportions for all metrics increase with increasing *k* unless there is high animal study heterogeneity, and decrease with increasing human study heterogeneity.

#### Assuming an animal null effect and a large human effect

(top right plot in Figure 2) Here, all metrics aside from the replication BF and the meta-analysis behave as one would expect: they rarely flag translation success. When there is no heterogeneity across animal studies, the proportion ranges from *α* for the two-trials rule to 10% for the golden sceptical *p*-value (high shrinkage) and the controlled sceptical *p*-value. These proportions further increase with increasing levels of animal study heterogeneity. Applying the strict criterion again results in proportions close to 1 for all metrics.

#### Summary

For a summary of the findings from our simulation study, we refer to Table 3.

## 4 Discussion

In our simulation study, we investigated whether metrics used or developed to assess replication success can be applied and are useful in the context of translation of results from animal studies to human studies. Our study was motivated by recorded cases of translation failure in biomedical research. We aimed to assess how well various metrics capture the concept of translation under a wide range of simulated conditions, including differences in effect sizes, effect size heterogeneity, animal study sample sizes and the number of animal studies pooled together. For this, we simulated animal and human studies using parameters informed by a real-world meta-analysis of prenatal amino acid supplementation on maternal blood pressure [28]. We also simulated different scenarios for the decision to move on to a human study: (1) any animal finding leads to a subsequent human study, (2) only beneficial animal findings lead to a subsequent human study, and (3) only significant beneficial animal findings lead to a subsequent human study. We evaluated nine metrics that have previously been discussed in the replication literature using pairs of findings from simulated animal and human studies.

We show that the performance of the different metrics highly depends on the simulation conditions. First, when both animal and human true effects are null, most metrics, except for the replication BF and meta-analysis, control the overall T1E rate close to the theoretical *α*^2^ under no heterogeneity. When heterogeneity increases, especially between human studies, the overall T1E increases. When both animals and humans had non-null effects, translation power was most influenced by whichever of the animal or human finding that had lower power. For example, under the conditions of small effects in both animals and humans and small animal sample sizes, translation power fell below (1 − *β*)^2^. Conversely, assuming large effects in both animals and humans yielded near-perfect translation power except in cases of high heterogeneity across animal studies, in which case translation power was lower.

Asymmetric effect size scenarios revealed systematic tendencies. Meta-analysis generally flagged success more often, driven by strong evidence in either animals or humans, while the two-trials rule and the golden sceptical *p*-value (no shrinkage) were more conservative and aligned more closely with the weaker of the two findings. Replication BF did not perform well whenever asymmetric effect sizes were simulated. These trends observed with asymmetric effect sizes could be explained by the fact that many replication success metrics treat findings as exchangeable and weight both studies equally, whereas in a translation scenario such a behaviour is not desired. Increasing the number of animal studies that were pooled (*k*) typically improved translation power when animal effects were non-null and heterogeneity was low, but had little benefit and even a negative impact when animal study heterogeneity was high, which is however often the case in practice.

Conditioning on significant animal findings as a strict criterion to move on to human testing, inflated animal effect size estimates and affected the operating characteristics of the metrics, e.g., sometimes substantially increasing T1E rates or power. Overall, no single metric was uniformly optimal. The controlled sceptical *p*-values and weighted Edgington performed relatively well across many scenarios, while replication BF and meta-analysis were highly sensitive to strong findings in either animals or humans. Golden sceptical *p*-values offered more conservative control at the cost of reduced power when true effects were small.

The replication and translation settings inherently differ. Replications follow the methodology of original studies closely and investigate the same research question in the same population, whereas translation investigates the same research question across two systematically different populations. Some replication success metrics were designed on the core assumption that original and replication studies investigate the same effect, and their performance in the translation context can, to some degree, be understood as a consequence of that assumption being violated.

A conceptual challenge uncovered in our simulation study was how to interpret cases in which the true effect sizes in animal and human differ in complex ways. For example, is a “translation success” desirable in the case where the true animal effect is null but the human effect is small? Most probably it is not. Such cases would benefit from a deeper discussion in the community of what constitutes a successful translation, especially because animal testing is often treated as a precursor to human studies rather than an end in itself. It is therefore important to recognize that translation differs fundamentally from replication, because in the translation setting the human finding is the reference point and the target population against which success is ultimately judged. Further, it is important to note that the metrics evaluated here should not never be viewed as stand-alone decision rules for progression from animal studies to human trials. Recent frameworks, such as PATH (Preclinical Assessment for Translation to Humans) [24] or the GALENOS approach [52, 25], show that translational decisions depend on a suite of evidence spanning mechanistic rationale, disease-model validity, pharmacology, safety, and efficacy. Within such a framework, translation success metrics may provide a quantitative assessment of the consistency between animal and human efficacy findings, thereby informing one component of a larger translational evidence assessment.

### Limitations

This study has various limitations. Our simulation study assumes a degree of comparability of effect sizes between animal and human studies that may not exist in practice. In reality, the magnitude of effects often differ substantially across species due to biological, methodological and environmental factors. The type of effects and outcome measurements investigated in animals might differ from those that are of interest in human studies. Human studies typically progress through various clinical phases with distinct goals and our simulation study did not distinguish between these phases.

While the parameters for the true animal and human study effect sizes and heterogeneity were based on a real-world motivating example from Terstappen et al. [28], we recognize that these values may not be representative in other biomedical contexts, especially the true animal effect size of −24.37mmHg and the animal study heterogeneity of 291.1mmHg. The human sample size used in our simulation study was fixed at *n*_H_ = 107 per group based on a power calculation for the specific question presented in Terstappen et al. [28]. We did not vary this sample size. Due to the fully factorial design, some combinations of simulation conditions may have also been unrealistic. However, the main goal of our study was not to determine the performance of the translation success metrics under specific values of effect sizes, heterogeneity, *etc*, but rather to analyse trends in the metrics’ behaviour as these values increased or decreased in general.

In addition, our choice to pool a maximum of five animal studies to form a single “animal finding” may be overly simplistic. In real-world settings, a larger number of animal studies might be required prior to deciding to continue to a human trial. Furthermore this decision is often not (solely) based on the statistical significance and direction of effect in the animal studies.

Our study solely investigated the statistical aspect of translation, which is only one component of a more complex decision-making process which involves consideration of other factors such as pharmacokinetics, safety profiles and ethical considerations. It is also important to emphasize that statistical translation is distinct from biological translation, which is primarily concerned with underlying mechanisms. Biological processes are extremely complex, involving the interplay of individual- and species-level differences in genetics and physiology, environmental factors, and disease mechanisms. This is reflected even in the motivating example in Terstappen et al. [28], in which different strains of rat used across studies may have contributed to heterogeneity. Furthermore, rat models used within a specific study are genetically similar and purpose-bred to investigate a specific condition, which does not reflect the diversity and comorbidities found in human populations. It would be inappropriate to reduce biological translation to a simple “yes or no” answer as done in statistical translation. We did not directly simulate publication bias of animal studies, which likely also contributes to the failure to translate because the decision to continue to human studies would rest upon incomplete information. Finally, we focused on a specific set of metrics, though other, more appropriate metrics might exist. Given those limitations, we invite investigators to explore other parameters and metrics using our openly available code.

### Recommendations

Our findings highlight that the choice of translation success metrics, along with the design features of both animal and human studies, can meaningfully influence conclusions about “translatability”. Generally, the low translation power of small-sample animal studies, even if effects are truly present, suggests that pooling multiple studies or increasing samples sizes is crucial to reduce false negatives and avoid inflated effect size estimates, especially when results will be used to justify human clinical trials. Special attention should also be given to heterogeneity when interpreting translation failures, as even modest heterogeneity across studies can reduce the chance of translation success according to most metrics. Our results also suggest caution when basing the justification of clinical trials solely on statistical significance in animal findings, *i.e*., the strict criterion, as this can lead to overly optimistic expectations for human outcomes. Recommendations for which translation success metrics to use are highly dependent on the expected animal and human study characteristics and the goal of the researcher in assessing translation success. For example, if the goal of translation is to infer that there is an effect in both animals and humans, we recommend using a metric which maintains good control of the overall T1E rate. Metrics such as the significance criterion, unweighted Edgington’s method, and the golden sceptical *p*-value (no shrinkage) perform well in this regard, though at the cost of reduced power. On the other hand, if the goal of translation is primarily to assess the evidence of an effect in humans, while still taking into account the animal results, then the researcher should use a metric which down-weights the evidence in the animal studies, *i.e*., weighted Edgington’s method. If high heterogeneity is anticipated in the animal and/or human studies, the performance of the significance criterion in terms of the overall T1E and translation power tends to be the least affected compared to the other metrics. Finally, if there are no strong assumptions with respect to heterogeneity or effect size, the controlled sceptical *p*-value tends to give consistent power across varying conditions, but is more conservative and leads to higher rates compared to others when there is a mismatch between true effect sizes.

In general, we advise against using the meta-analysis metric or replication BF to assess translation success. The meta-analysis metric tends to have the highest translation power, at the cost of the highest overall T1E rate. This is due to the fact that it will often flag success, even if only one of the two studies shows an effect. However, if the researcher wishes to assess the evidence for a combined/pooled effect, then the meta-analysis metric can be appropriate. The performance of the replication BF tends to perform poorly across most conditions in terms of overall power, except in the case of high animal or human heterogeneity in which it becomes comparable to the other metrics.

Ultimately, no one metric performed best across all simulation conditions. We recommend using a combination of metrics which emphasize different aspects of translation success and making a decision based on the combined result. In general, metrics that balance information from both animals and humans – such as controlled sceptical *p*-values or weighted Edgington – may provide more robust conclusions than metrics that are driven by strong evidence in just one species (*e.g*., replication BF which focuses mainly on human findings, and meta-analysis).

## Conclusions

We conclude that metrics developed for assessing replication success can offer valuable insights for assessing translation success. However, their utility depends strongly on the context, underlying assumptions, and the characteristics of the available evidence. No single metric performed optimally across all simulated scenarios. A combined approach, using multiple metrics alongside an understanding of their respective strengths and limitations, is recommended to assess when and how animal findings translate to human outcomes. Future research is needed to explore and better understand the behaviour of the metrics in the translation setting from a theoretical perspective to draw generalisable conclusions in biomedical contexts.

## Data Availability

All data and code file to reproduce our simulation results, this manuscript and the online supplement are available via GitLab, https://gitlab.uzh.ch/rachelheyard/translation_simulation. A citable snapshot of the repository at the time writing is archived at https://doi.org/10.5281/zenodo.13587432.

https://rachelheyard.pages.uzh.ch/translation_simulation/

https://gitlab.uzh.ch/rachelheyard/translation_simulation

## Data and software availability

All data and code file to reproduce our simulation results, this manuscript and the online appendix are available via GitLab, https://gitlab.uzh.ch/rachelheyard/translation_simulation. A citable snapshot of the repository at the time writing is archived at https://doi.org/10.5281/zenodo.13587432.

## Acknowledgements

We thank Gillian Currie and Bernhard Voelkl for valuable feedback on an earlier version of our manuscript. Additionally, we would like to thank the iRISE consortium, and specially work package 1, for continuous feedback in the conceptualization and reporting of our work.

## Author contributions

1. **Conceptualization:** CJH, SP, KEW, BVI, RH
2. **Data curation:** CJH, KEW
3. **Formal Analysis:** CJH, RH
4. **Funding acquisition:** KEW, BVI, RH
5. **Methodology:** CJH, SP, RH
6. **Project administration:** RH
7. **Software:** CJH, RH
8. **Supervision:** RH
9. **Visualization:** CJH, RH
10. **Writing – original draft:** CJH, RH
11. **Writing – review & editing:** CJH, RH SP, KEW, BVI

## Appendix

### Complete Figures

Figure 1.A shows the results of the simulation study for all metrics, also replication BF and meta-analysis, across scenarios when the animal studies’ sample size is fixed to 10 per group. Figure 2.A shows the same type of results for when the animal studies’ sample size is fixed to 20 per group, while Figure 3.A shows the zoomed-in results where replication BF and meta-analysis were dropped for readability.

**Figure A.1:**
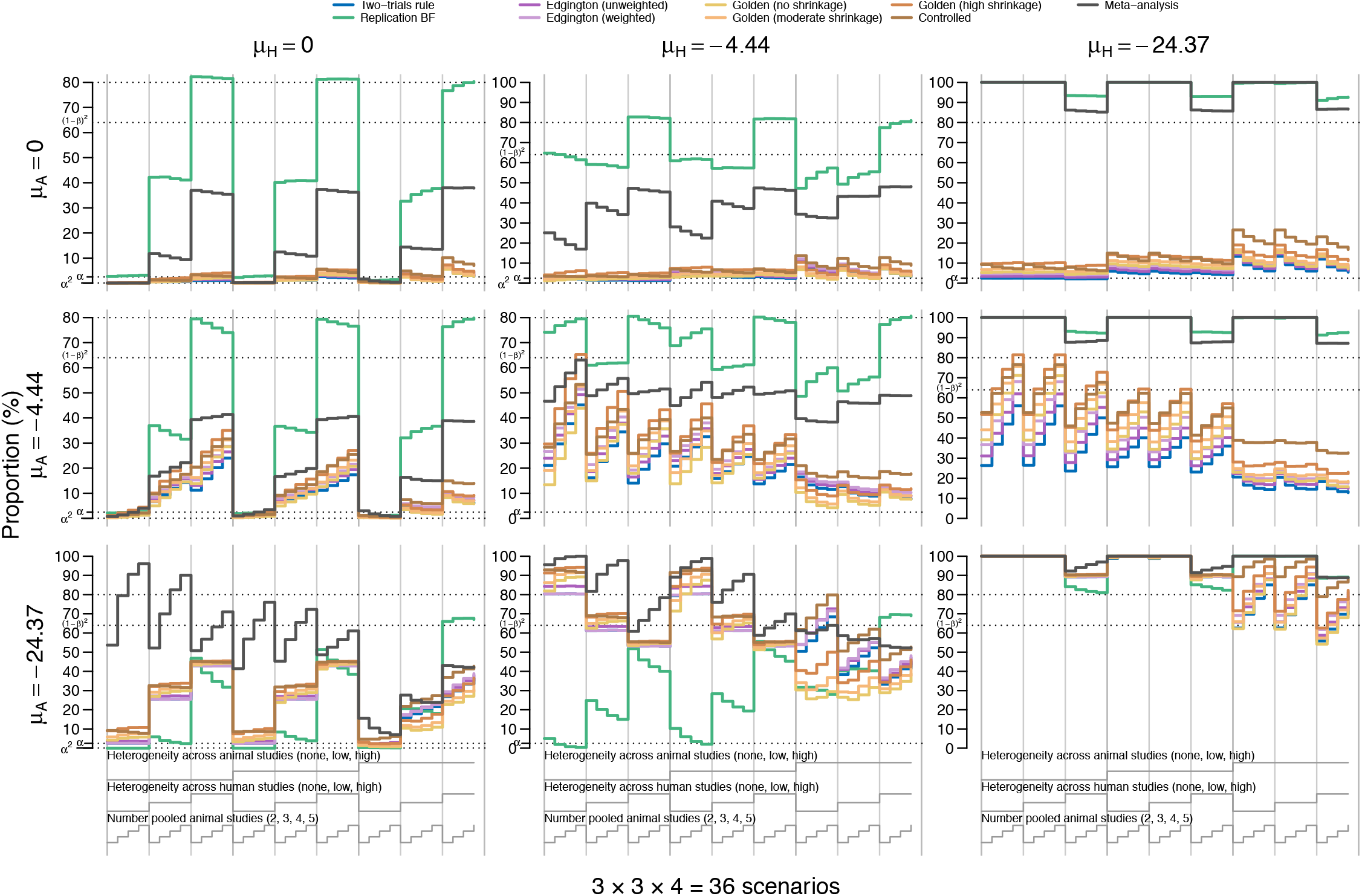
Grid of nested loop plots of the proportions of animal-human pairs for which the different metrics flagged successful translation across simulation conditions under no criterion. Each of the plots in the grid represent another animal-human finding combination. In the first column, for example, the human studies are all simulated under the null hypothesis of no effect. The dotted horizontal lines represent *α*^2^ = 0.000625, *α* = 0.025, 1 − β = 0.8 and (1 − β)^2^ = 0.64. All animal studies in this representation are simulated with a small sample size per group (n_A_ = 10).

**Figure A.2:**
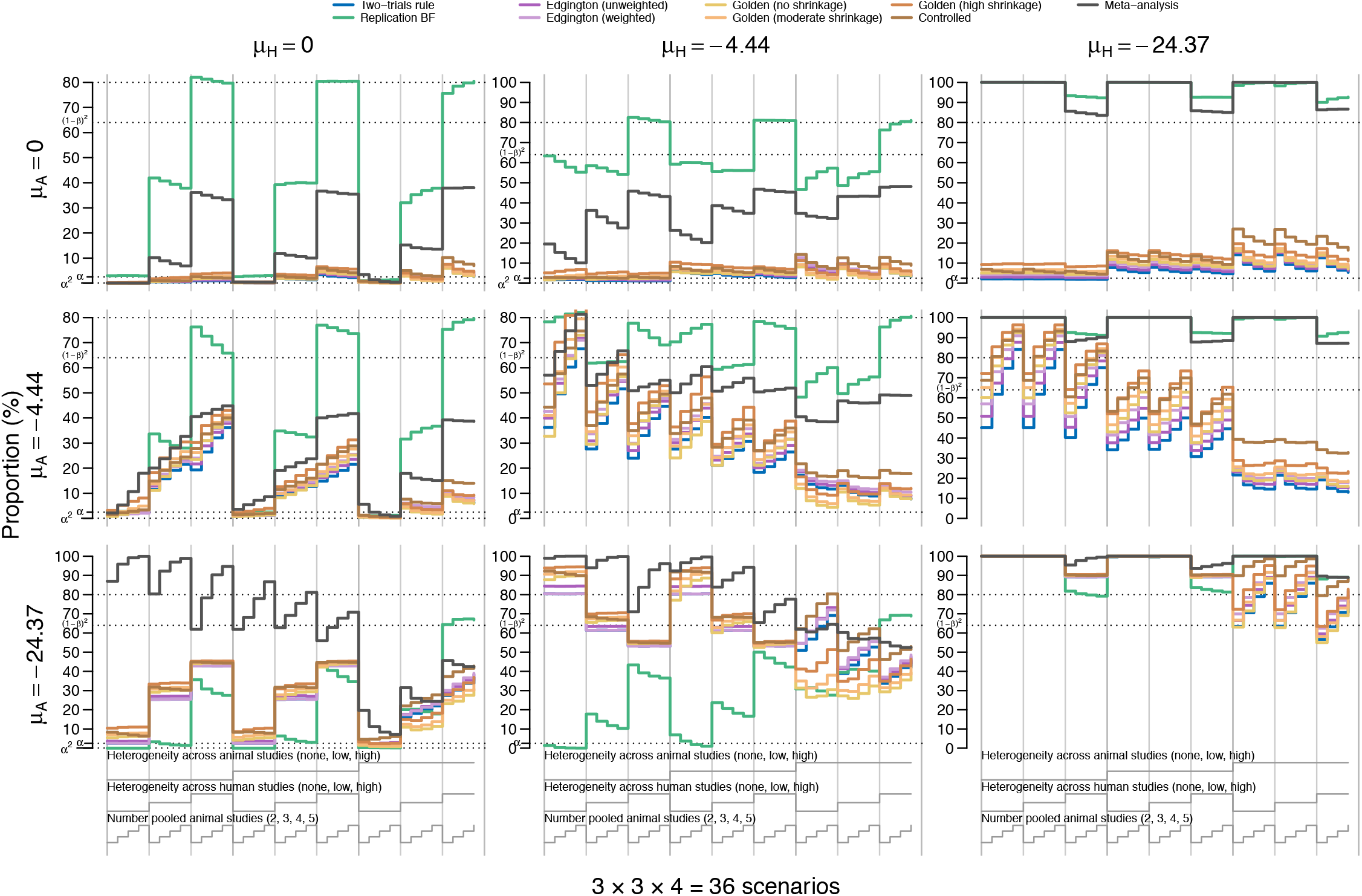
Grid of nested loop plots of the proportions of animal-human pairs for which the different metrics flagged successful translation across simulation conditions under no criterion. Each of the plots in the grid represent another animal-human finding combination. In the first column, for example, the human studies are all simulated under the null hypothesis of no effect. The dotted horizontal lines represent α^2^ = 0.000625, α = 0.025, 1 − β = 0.8 and (1 − β)^2^ = 0.64. All animal studies in this representation are simulated with a larger sample size per group (n_A_ = 20).

**Figure A.3:**
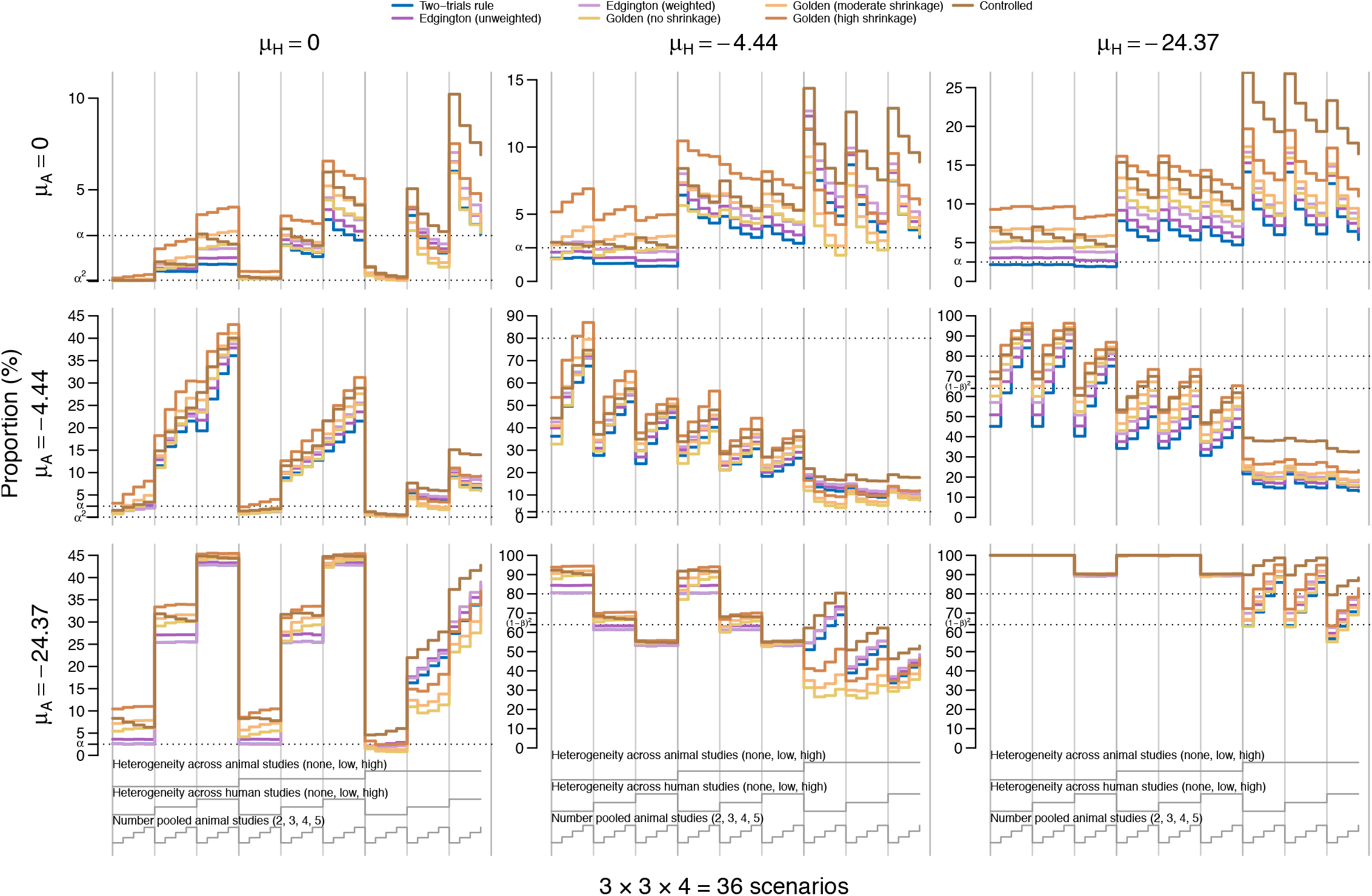
Grid of nested loop plots of the proportions of animal-human pairs for which the different metrics flagged successful translation across simulation conditions under no criterion. Each of the plots in the grid represent another animal-human finding combination. In the first column, for example, the human studies are all simulated under the null hypothesis of no effect. Note that the results for the replication BF and the meta-analysis are not shown here for better readability. The dotted horizontal lines represent *α*^2^ = 0.000625, *α* = 0.025, 1 − *β* = 0.8 and (1 − *β*)^2^ = 0.64. All animal studies in this representation are simulated with a larger sample size per group (*n*_A_ = 20).

## Footnotes

1 While this upper value may appear high, such effect sizes, on the raw mean difference scale, are commonly reported in animal research [29], and including extreme cases is important for a thorough evaluation of metric performance

2 We refer to the caption of Figure 1.(a) for a brief description of how to interpret these plots.

## References

[1] B. V. Ineichen, E. Furrer, S. L. Grüninger, W. E. Zürrer, and M. R. Macleod. “Analysis of animal-to-human translation shows that only 5% of animal-tested therapeutic interventions obtain regulatory approval for human applications”. In: PLoS Biology 22.6 (2024), e3002667.

[2] C. H. Leenaars et al. “Animal to human translation: A systematic scoping review of reported concordance rates”. In: Journal of translational medicine 17 (2019), pp. 1–22.

[3] D. G. Hackam and D. A. Redelmeier. “Translation of research evidence from animals to humans”. In: Jama 296.14 (2006), pp. 1727–1732.

[4] S. Perrin. “Preclinical research: Make mouse studies work”. In: Nature 507.7493 (2014), pp. 423–425.

[5] B. Voelkl et al. iRISE Reproducibility Glossary. 2024. doi: 10.17605/OSF.IO/BR9SP.

[6] A. Schmidt-Pogoda et al. “Why most acute stroke studies are positive in animals but not in patients: a systematic comparison of preclinical, early phase, and phase 3 clinical trials of neuroprotective agents”. In: Annals of neurology 87.1 (2020), pp. 40–51.

[7] E. Wilson et al. “Designing, conducting, and reporting reproducible animal experiments”. In: Journal of Endocrinology 258.1 (2023).

[8] S. C. Landis et al. “A call for transparent reporting to optimize the predictive value of preclinical research”. In: Nature 490.7419 (2012), pp. 187–191.

[9] J. D. Wallach, K. W. Boyack, and J. P. A. Ioannidis. “Reproducible research practices, transparency, and open access data in the biomedical literature, 2015–2017”. In: PLOS Biology 16.11 (2018). Ed. by U. Dirnagl, e2006930. doi: 10.1371/journal.pbio.2006930.

[10] K. S. Button et al. “Power failure: why small sample size undermines the reliability of neuroscience”. In: Nature Reviews Neuroscience 14.5 (2013), 365–376. doi: 10.1038/nrn3475.

[11] A. Bespalov et al. “Failed trials for central nervous system disorders do not necessarily invalidate preclinical models and drug targets”. In: Nature Reviews Drug Discovery 15.7 (2016), pp. 516–516.

[12] R.-D. Gosselin. “Insufficient transparency of statistical reporting in preclinical research: a scoping review”. In: Scientific Reports 11.1 (2021). doi: 10.1038/s41598-021-83006-5.

[13] C. Kilkenny et al. “Survey of the quality of experimental design, statistical analysis and reporting of research using animals”. In: PLoS ONE 4.11 (2009), e7824.

[14] E. S. Sena, H. B. Van Der Worp, P. M. Bath, D. W. Howells, and M. R. Macleod. “Publication bias in reports of animal stroke studies leads to major overstatement of efficacy”. In: PLoS Biology 8.3 (2010), e1000344.

[15] D. Fanelli. “Negative results are disappearing from most disciplines and countries”. In: Scientometrics 90.3 (2012), pp. 891–904.

[16] P. Flecknell. “Replacement, reduction, refinement”. In: ALTEX-Alternatives to animal experimentation 19.2 (2002), pp. 73–78.

[17] National Academies of Sciences, Engineering, and Medicine. Reproducibility and Replicability in Science. National Academies Press, 2019. doi: 10.17226/25303.

[18] R. Heyard et al. “A scoping review on metrics to quantify reproducibility: a multitude of questions leads to a multitude of metrics”. In: Royal Society Open Science 12.7 (2025). doi: 10.1098/rsos.242076.

[19] F. Freuli, L. Held, and R. Heyard. “Replication success under questionable research practices—a simulation study”. In: Statistical Science 38.4 (2023), pp. 621–639.

[20] J. Muradchanian, R. Hoekstra, H. Kiers, and D. van Ravenzwaaij. “How best to quantify replication success? A simulation study on the comparison of replication success metrics”. In: Royal Society Open Science 8.5 (2021), p. 201697.

[21] J. Muradchanian, R. Hoekstra, H. Kiers, and D. van Ravenzwaaij. “Evaluating meta-analysis as a replication success measure”. In: PLoS ONE 19.12 (2024). Ed. by D. Purić, e0308495. doi: 10.1371/journal.pone.0308495.

[22] L. Held. “Beyond the two-trials rule”. In: Statistics in Medicine (2024).

[23] B. A. Nosek and T. M. Errington. “Making sense of replications”. In: eLife 6 (2017). doi: 10.7554/elife.23383.

[24] J. Kimmelman, P. Bodilly Kane, S. Bicer, and B. G. Carlisle. “Preclinical assessment for translation to humans: The PATH approach for assessing supporting evidence for early-phase trials and innovative care”. In: Med 5.10 (2024), 1227–1236. doi: 10.1016/j.medj.2024.07.014.

[25] T. Tonia et al. “The GALENOS approach to triangulating evidence: a structured approach for integrating information from human and animal studies”. In: BMC Medical Research Methodology (2026). doi: 10.1186/s12874-026-02891-4.

[26] B. S. Siepe et al. “Simulation studies for methodological research in psychology: A standardized template for planning, preregistration, and reporting.” In: Psychological Methods (2024). doi: 10.1037/met0000695.

[27] C. J. Huang and R. Heyard. A simulation study to quantify successful translation of results from preclinical studies to human trials. 2024. doi: 10.17605/OSF.IO/BZXVY.

[28] F. Terstappen et al. “Prenatal amino acid supplementation to improve fetal growth: a systematic review and meta-analysis”. In: Nutrients 12.9 (2020), p. 2535.

[29] K. E. Wever et al. “Determinants of the Efficacy of Cardiac Ischemic Preconditioning: A Systematic Review and Meta-Analysis of Animal Studies”. In: PLOS ONE 10.11 (2015). Ed. by T. Eckle, e0142021. doi: 10.1371/journal.pone.0142021.

[30] C. R. Hooijmans et al. “Assessment of key characteristics, methodology, and effect size measures used in meta-analysis of human-health-related animal studies”. In: Research Synthesis Methods 13.6 (2022), 790–806. doi: 10.1002/jrsm.1578.

[31] C. R. Hooijmans et al. “Remyelination promoting therapies in multiple sclerosis animal models: a systematic review and meta-analysis”. In: Scientific Reports 9.1 (2019). doi: 10.1038/s41598-018-35734-4.

[32] J. Y. Chien, S. Friedrich, M. A. Heathman, D. P. de Alwis, and V. Sinha. “Pharmacokinetics/pharmacodynamics and the stages of drug development: role of modeling and simulation”. In: The AAPS Journal 7 (2005), E544–E559.

[33] P. C. Lind et al. “Translation from animal studies of novel pharmacological therapies to clinical trials in cardiac arrest: A systematic review”. In: Resuscitation 158 (2021), pp. 258–269.

[34] G. K. Rosenkranz. “A Generalization of the Two Trials Paradigm”. In: Therapeutic Innovation & Regulatory Science 57.2 (2022), 316–320. doi: 10.1007/s43441-022-00471-4.

[35] R. D. Cousins. Annotated Bibliography of Some Papers on Combining Significances or p-values. 2007. doi: 10.48550/ARXIV.0705.2209.

[36] C. Röver and T. Friede. “Investigating the Heterogeneity of “Study Twins”“. In: Biometrical Journal 66.6 (2024). doi: 10.1002/bimj.202300387.

[37] J. Verhagen and E.-J. Wagenmakers. “Bayesian tests to quantify the result of a replication attempt.” In: Journal of Experimental Psychology: General 143.4 (2014), p. 1457.

[38] H. Jeffreys. The theory of probability. 3rd ed. Oxford Classic Texts in the Physical Sciences. London, England: Oxford University Press, 1998.

[39] E. S. Edgington. “An additive method for combining probability values from independent experiments”. In: The Journal of Psychology 80.2 (1972), pp. 351–363.

[40] L. Held, S. Pawel, and C. Micheloud. “The assessment of replicability using the sum of p-values”. In: Royal Society Open Science (2024). doi: 10.48550/arXiv.2401.13615.

[41] L. Held. “A new standard for the analysis and design of replication studies”. In: Journal of the Royal Statistical Society Series A: Statistics in Society 183.2 (2020), pp. 431–448.

[42] L. Held, C. Micheloud, and S. Pawel. “The assessment of replication success based on relative effect size”. In: The Annals of Applied Statistics 16.2 (2022), pp. 706–720.

[43] C. Micheloud, F. Balabdaoui, and L. Held. “Assessing replicability with the sceptical p-value: Type-I error control and sample size planning”. In: Statistica Neerlandica 77.4 (2023), pp. 573–591.

[44] Open Science Collaboration. “Estimating the reproducibility of psychological science”. In: Science 349.6251 (2015). doi: 10.1126/science.aac4716.

[45] T. M. Errington et al. “Investigating the replicability of preclinical cancer biology”. In: eLife 10 (2021). doi: 10.7554/elife.71601.

[46] G. Rücker and G. Schwarzer. “Presenting simulation results in a nested loop plot”. In: BMC Medical Research Methodology 14.1 (2014). doi: 10.1186/1471-2288-14-129.

[47] T. P. Morris, I. R. White, and M. J. Crowther. “Using simulation studies to evaluate statistical methods”. In: Statistics in Medicine 38.11 (2019), 2074–2102. doi: 10.1002/sim.8086.

[48] R. P. Chalmers and M. C. Adkins. “Writing effective and reliable Monte Carlo simulations with the SimDesign package”. In: The Quantitative Methods for Psychology 16.4 (2020), pp. 248–280. doi: 10.20982/tqmp.16.4.p248.

[49] S. Pawel and L. Held. The sceptical Bayes factor for the assessment of replication success. 2022. doi: 10.1111/rssb.12491.

[50] L. Held, C. Micheloud, S. Pawel, F. Gerber, and F. Hofmann. Design and Analysis of Replication Studies with ReplicationSuccess. 2022. doi: 10.32614/CRAN.package.ReplicationSuccess.

[51] S. Pawel, F. Bartoš, B. S. Siepe, and A. Lohmann. “Handling Missingness, Failures, and Non-Convergence in Simulation Studies: A Review of Current Practices and Recommendations”. In: The American Statistician 80.1 (2026), pp. 31–48. doi: 10.1080/00031305.2025.2540002.

[52] K. A. Smith et al. “Triangulating evidence from the GALENOS living systematic review on trace amine-associated receptor 1 (TAAR1) agonists in psychosis”. In: The British Journal of Psychiatry 226.3 (2024), 162–170. doi: 10.1192/bjp.2024.237.

